# Characterization of Clinical MRI Findings in Moderately-Late Preterm Infants Diagnosed with Cerebral Palsy: A Single Center Retrospective Study

**DOI:** 10.1101/2024.09.20.24313922

**Authors:** Elizabeth Fisher, Jessica Tartakovsky, Laura A. Bliss, Alok Jaju, Jessie Aw-Zoretic, Laura Vernon, Divakar S. Mithal

## Abstract

Cerebral Palsy is the most common movement disorder in childhood and is commonly associated with brain injury and prematurity. At least 10% of patients have a normal brain MRI and current practices suggest genetic testing may be indicated for those patients. However, given that prematurity itself is a risk factor for CP, which MRI patterns are present in premature infants and which genetic causes are present remains unclear. In the present study, a cohort of moderate-late preterm infants born between 32 and 34 weeks gestational age was analyzed to determine whether CP disease severity, comorbid conditions, and genetic testing are associated with particular MRI findings. We identified that comorbid conditions and CP severity are similar across imaging patterns regardless of the perceived severity of the imaging findings. Genetic diagnoses were identified in 13% of all patients. In patients with MRI concerning for injuries, over 20% had genetic testing sent. Abnormal MRI findings in moderate-late preterm infants with CP are common, but are not a strong predictor of disease severity and may be associated with underlying genetic causes that are underdiagnosed due to unclear practice guidelines.

## II. Introduction

Cerebral palsy (CP) is the most common movement disorder diagnosed in childhood, presenting in every 2-3 per 1,000 births^1^. CP is a clinical syndrome characterized by developmental disorders of movement and posture attributed to an injury or insult to the developing fetal or infant brain, often accompanied by muskoluskeletal and seizure disorders as well as impaired sensation, cognition, and communication ^2^. Though the majority of patients with CP are born at or near term, prematurity is one of the few well-defined risk factors for CP^3^. In infants born at term, seizures in the neonatal period, birth asphyxia, and major and minor birth defects have been identified as major risk factors for CP^4^. However, the etiology of CP specifically in preterm infants is not well characterized. Understanding the etiology of CP may help understand the overall prognosis, as children with CP are at increased risk for multiple comorbidities, including epilepsy, intellectual disability, digestive diseases, and respiratory infections, in relation to their underlying CP^5^.

International experts agree that CP is secondary to some abnormality or lesion in the developing brain, and as such MRI is the first line tool for diagnosing CP in children, demonstrating a sensitivity of 86-89% in infants less than 12 months of age^6,7^. Previous work has demonstrated that MRI findings may differ between children born before 34 weeks gestational age (WGA) versus children born after 34 WGA^8^. However, it is not yet known whether the severity of brain MRI findings correlates withCP functional status or global disease burden across gestational ages. Furthermore, recent data indicates that 10.5% of children with CP may have an unremarkable brain MRI, and in such cases genetic testing is a logical next step in diagnosis^9^.

With the increasing utility of genetic testing, the role of genetic variants in CP etiology is becoming increasingly apparent. Two meta-analyses indicate that genetic variants may be present from 23% to 31% of individuals with CP^10,11^. However, the extent to which genetic variants play a role in CP still remains controversial. One study with 151 participants found a genetic etiology in 8% of patients^12^, yet another study of 150 patients with CP demonstrated pathogenic or likely pathogenic variants in 24.7% of participants^13^. These studies did not take gestational age into account, so while the promise of genetic testing is now an active field of interest for the CP community, the role of genetic testing specifically for premature infants remains unclear^14^.

The work presented in this study seeks to: 1) characterize brain MR findings in a cohort of individuals with CP born moderately preterm between the gestational ages of 32 weeks and 33 weeks 6 days, at a single research hospital; 2) identify any correlation between brain MRI findings and genetic testing patterns and; 3) characterize the ambulatory status, verbal status, and presence of intellectual disability in patients by MRI imaging group to determine the utility of MRI as a marker of CP functional status; and 4)identify trends in prevalence of epilepsy, tracheostomy, and gastrostomy tube in each MRI imaging group as a marker of global disease burden.

## III. Patients and Methods

The study received Institutional Review Board approval from Ann & Robert H Lurie Children’s Hospital of Chicago. A retrospective cohort study was performed on infants born at a single tertiary care children’s hospital between 2007 and 2020 with CP ICD codes. Patients without a brain MRI report available for review were excluded from the study. The included patients born between 32w0d and 33w6d with MRI brain imaging reports were grouped into five categories based on a predetermined set of criteria defining the likelihood that MRI findings were causative of a patient’s CP: 1) Normal, 2) Nonspecific, Unlikely Causal, 3) Nonspecific, Likely Causal 4) Acquired Pathology, and 5) Congenital/ Structural. MRI categorization was based entirely on the written imaging findings of the radiologist at the time of imaging, and assignment to a category was completed by consensus amongst the authors using criteria that can be found in Table 1. MRI imaging was not directly visualized in this study. In addition to MRI data, the patient’s sex and birth weight were recorded to identify potential confounding variables for demographic purposes. The patient history of genetic testing and any genetic diagnoses were recorded.

**Table 1:**
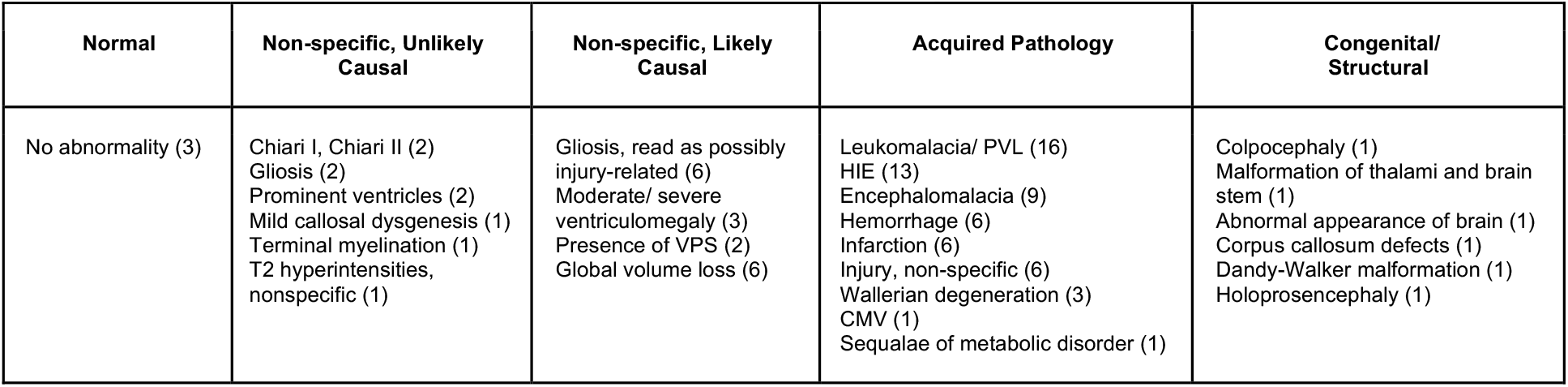
Criteria for MRI Categorization. Patients were categorized into one of five groups based off the most severe finding in their earliest Neuroradiology report. The numbers in parentheses indicated the number of patients with that particular imaging finding. 96% of patients in this sample had some remarkable finding on MRI. The most common finding among all patients was leukomalacia/periventricular leukomalacia (PVL) followed by hypoxic-ischemic encephalopathy (HIE)

Tracheostomy tube dependence, gastrostomy tube dependence, and epilepsy or seizure diagnosis were recorded as markers of global disease burden. Markers of CP severity including ambulatory status, verbal status, and intellectual disability were recorded; the authors investigating markers of CP severity were blinded to the MRI categorization of each patient. Statistical analysis was conducted using OpenEpi Version 3 software. Fisher’s exact test was used for analysis of the rate of genetic testing, the rate of genetic diagnosis, tracheostomy tube dependence, gastrostomy tube dependence, epilepsy or seizure diagnosis, ambulatory status, verbal status, and intellectual disability. A two-tailed t-test was used to compare the average weights across each MRI sub-cateogry.

## IV. Results

Our criteria yielded 104 patients born between 32w0d and 33w6d with CP. Among these patients, 65 patients (62%) had MRI-Brain imaging reports available for review and were included in the study cohort. In the entire cohort, 95% had at least one finding in their MRI report. Acquired Pathology was apparent in 54%. Additionally, 33% of patients had Non-specific imaging findings: 20% of patients were Non-specific, Likely Causal, while 12% were Non-specific, Unlikely Causal. A Congenital/Structural abnormality accounted for 9% of patients. Finally, only 3 patients (5%) had MRI imaging that was read as completely normal (Figure 1). Among patients with Acquired Pathology, the most common finding was leukomalacia/periventricular leukomalacia (PVL), which was present in 46% of the MRIs in this group. Hypoxic-ischemic encephalopathy (HIE) was the next most common finding (37%) followed by encephalomalacia (26%). Hemorrhage, infarction, and non-specific injury occurred with equal frequency (17%). Wallerian degeneration, infection, and metabolic disorders were less common. Many of the Acquired Pathology MRI had multiple injury-related findings (data not shown). Notably, within the cohort, 46% were female with no statistically significant sex differences across the imaging groups (Table 2). Additionally birthweight was determined for all included patients, and the only group-by-group difference identified was between the Congenital/Structural and Non-specific Likely Causal group (Table 2).

**Table 2.** Image Group Comparisons. There was no statistical difference in the distribution of females in each MRI group. There was a statistically significant difference in weight between the weight of the Congenital/Structural and Non-specific, Likely causal groups by t-test (p=0.0329).

| MRI Categorization, n | Female, n (%) | Average weight in grams (SD) |
| --- | --- | --- |
| Normal, 3 | 1 (33) | n/a |
| Congenital/ Structural, 6 | 4 (67) | 2174 (580)* |
| Non-specific, Unlikely Causal, 8 | 3 (38) | 1655 (529) |
| Non-specific, Likely Causal, 13 | 4 (31) | 1482 (473)* |
| Acquired Pathology, 35 | 17 (49) | 2073 (866) |

**Figure 1.**
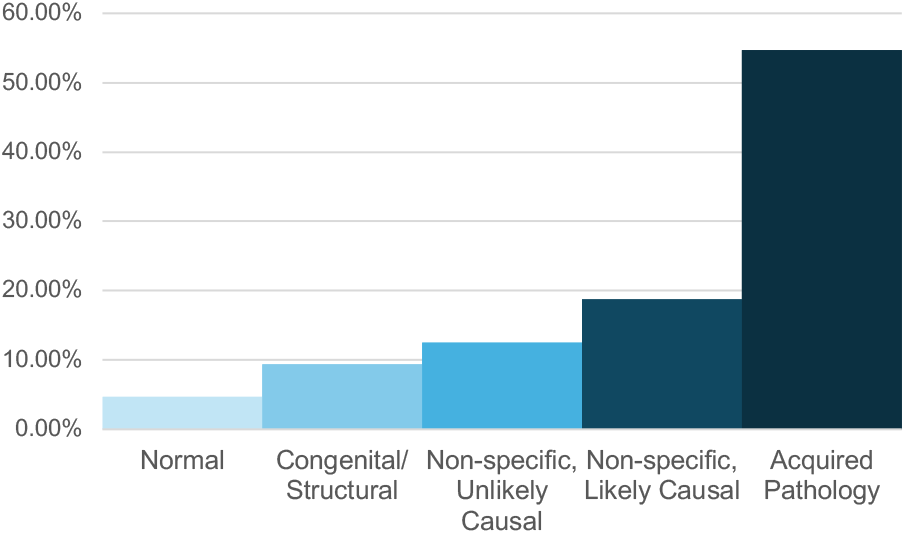
Percentage of patients from the cohort in each imaging group.

In the cohort, 34% of all patients received genetic testing, with a genetic diagnosis found in 13% of all patients, for a diagnostic yield of 34% (Figure 2). The Normal group had a genetic testing rate of 67% with a diagnostic yield of 50%. Genetic testing was most frequently sent in the Non-specific, Unlikely Causal group (75%). The Congenital/Structural group had the highest diagnostic yield at 67%. The Non-specific Likely Causal group had genetic testing in 33% of patients with a 25% diagnostic yield. The Acquired Pathology group received the least amount of genetic testing (20%) and had a diagnostic yield of 29%. Although the rates varied considerably across groups, there was no statistically significant difference by Fisher’s exact test in the rate of genetic testing or diagnostic yield across all five MRI cohorts (Figure 2).

**Figure 2.**
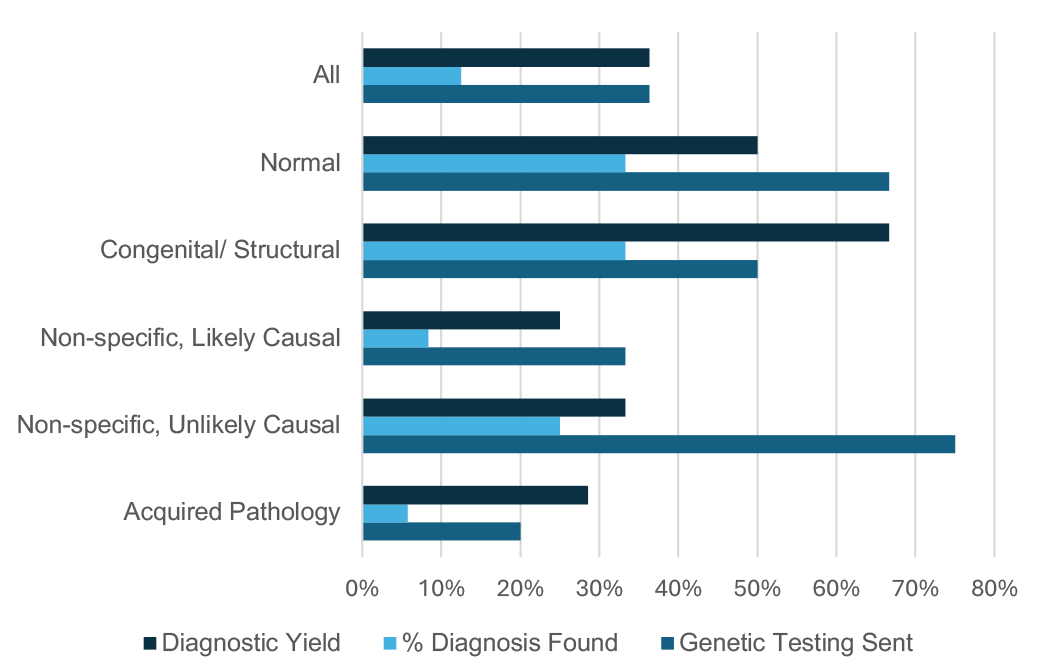
Testing rate, diagnostic rate and diagnostic yield for genetic testing in a moderately premature cohort of children with CP. Blue bars represent the Percent of patients for whom any genetic testing was sent. The light blue bars represent the percent of patients for whom a genetic diagnosis was found. The dark blue bar represents the resultant diagnostic yield of genetic testing. None of the differences across groups amounted to statistical significance.

Across the entire cohort, 64% of patients had an epilepsy/seizure diagnosis, 41% had gastrostomy tube, and 14% had tracheostomy tube (Figure 3). The rates of clinical comorbidities followed a similar pattern across the different MRI groups, with Epilepsy/seizure diagnosis was the highest comorbidity, followed by G tube dependence and Tracheostomy dependence. The clinical features across the different imaging groups were not statistically different.

**Figure 3.**
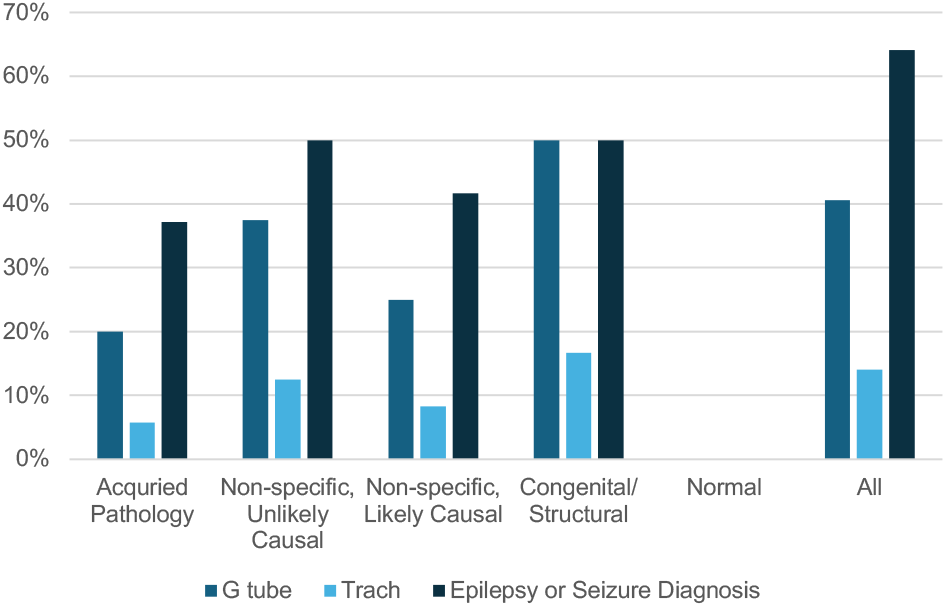
Clinical Co-morbidity in CP across different imaging groups. There was no statistical difference between the rate of epilepsy/seizure diagnosis, gastrostomy tube, or tracheostomy tube among all five groups, though none of the three comorbidities were present in the Normal group.

In evaluating CP severity, in our complete cohort, 36% of all patients were non-verbal, 34% were non-ambulatory, and 44% had intellectual disability. There was no statistically significant difference in the rate of non-verbal status, non-ambulatory status, and intellectual disability across all five MRI cohorts. Non-specific, Unlikely Causal group had the highest percentage of intellectual disability (71%), followed by the Congenital/Structural group (68%) (Figure 4).

**Figure 4.**
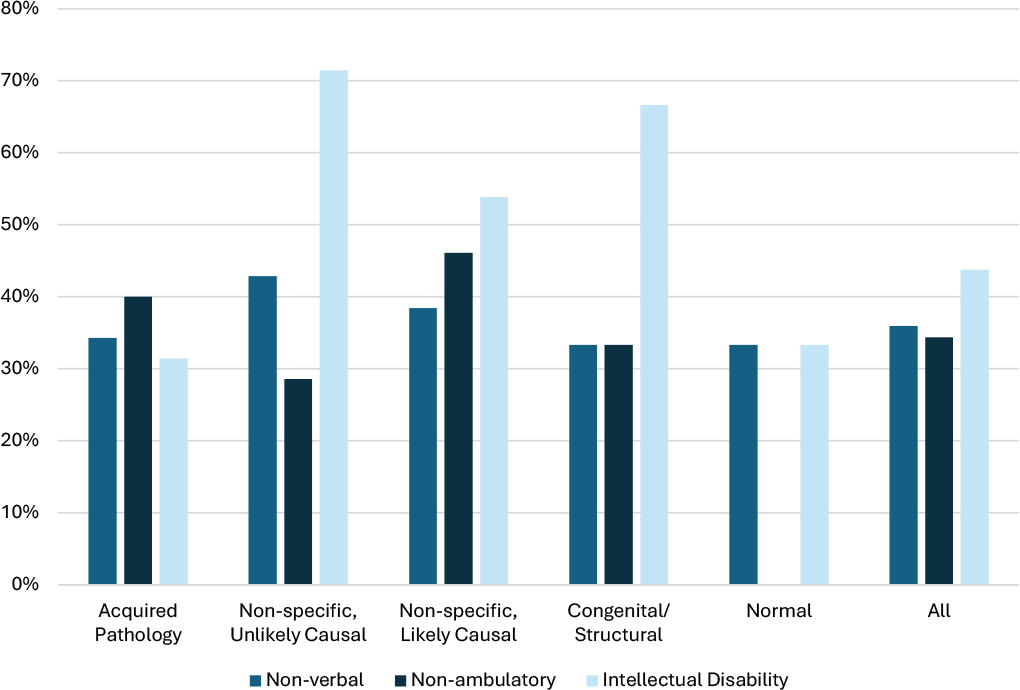
CP clinical severity across imaging groups. The light blue bars represent percent of patients with intellectual disability. The dark blue bars represent percent of patients who are non-ambulatory. The blue bars represent percent of patients who are non-verbal. All groups imaging groups were similar to the overall cohort. There was no statistically difference between imaging groups for any parameter.

## V. Discussion

In the present study, imaging patterns in late pre-term infants with CP were evaluated for etiology, disease comorbidity and CP severity. Extensive studies have been done of the nature of brain injuries as they are related to CP, but our study is one of the few to focus on the late-preterm gestational age. A total of 54% of our patients had MRI findings indicative of an Acquired Pathology, while an additional 19% of patients had Non-specific, Likely Causal findings that are strongly indicative of an injury pattern that caused the patient’s CP. These findings are more severe than those in the general pediatric population born in the moderate and late preterm stage. In a prospective study of all infants born between 32 and 36 WGA, only 3.6% of infants demonstrated moderate-severe lesions, which included IVH Grade III, post-hemorrhagic ventricular dilatation, cystic white matter lesions, periventricular hemorrhagic infarction, arterial infarction, and ex-vacuo ventricular dilatation^15^. Brain injury on MRI is the gold standard for diagnosing CP, indicating that the higher percentage of patients in our cohort with severe injury is likely consistent with our understanding of CP ^6^. However, the distribution of MRI findings in our cohort may also differ from the general CP population. Various sources report around 85% of patients have some kind of abnormality, while around 11% have normal imaging^16,17^. In this late pre-term group, 95% of patients had some kind of abnormality on brain MRI. While prematurity is a well-defined risk factor for CP, the majority of patients with CP are born at term, and would represent the dominant subtype within a general CP population^3^. When evaluating our premature infants and comparing our sample to the non-CP pre-term MRIs, it suggests that abnormal MRI patterns in moderately pre-mature infants are closely linked to CP^15^. Yet, the signifance and etiology of abnormal MRI findings in moderate preterm infants remains unclear and is an active area of further exploration.

Although brain injuries are linked to CP, our study did not demonstrate a statically significant difference in CP severity or global disease burden markers between any of the MRI cohorts, including those with a defined brain injury. In contrast, a previous, larger study of infants with CP with no exclusion criteria based on gestational age found that type of brain injury strongly correlated with severity of clinical presentation^8^. However, our sample exclusively included patients born between 32w0d and 33w6d, while only 18.3% of the larger study sample was born between 32 and 36 WGA, with the majority (54.5%) of patients being born at term^8^. This discrepancy between MRI findings and CP severity could be explained by our smaller sample size, or by our exclusive focus on patients born moderately-prematurely. Compared to another study which did not stratify patients by gestational age which reported 22% of patients were non-verbal, 36% of our cohort was non-verbal. This study also reported 16.9% of patients having an afebrile seizure disorder, while 64% of our sample had epilepsy or an alternative seizure diagnosis. The authors also reported 33% of patients being non-ambulatory by GMFCS criteria, and our cohort was similar^18^. As we selected for patients with MRI reports, it may be possible our patients born between 32w0d and 33w6d experienced a more severe disease course compared to the general CP population which could enrich for abnormal MRI findings. However, the Non-specific unlikely group, who had the mildest MRI findings in our cohort, had similar disease comorbidity and CP severity with the other groups, suggesting that MRI findings alone do not account for the CP disease process in our cohort.

To evaluate additional etiologies of CP, we examined genetic testing rates. Genetic testing was utilized in 34% of patients, the majority of which had Normal or Non-specific, Unlikely Causal MRI findings. However, few diagnoses were identified and there was no statistically significant difference between the rates or yield of genetic testing in any MRI sub-cohort. Notably, although the percent of patient tested in the Non-specific Unlikely Causal group was nearly double that of the Non-specific likely causal and Acquired Pathology groups, the diagnostic yield was similar. This findings suggests that perhaps genetic was underutilized in the injury-associated groups. An additional possibility is that the perceived injuries on the MRI were actually sequela of an underlying genetic disorder and not due to perinatal distress.

The utility of genetic testing in patients with CP is an emerging field, and there are no clear guidelines for selecting patients who would most likely benefit from genetic testing. A large meta-analysis found that the diagnostic yield of whole exome sequencing in patients with cryptogenic CP (or, CP without a common risk factor such as prematurity or birth asphyxia) was 35% compared to 7% in patients with known risk factors for CP, suggesting that genetic testing may be better utilized in patients without a presumed cause of their CP^10^. In our cohort, the imaging findings did not appear stratify genetic risk, but the rates of genetic testing were low across the entire cohort, making it difficult to conclude how efficacious testing could be.

Given the similarity in disease severity across imaging groups, genetic testing as an additional component of Cerebral Palsy workup may have utility. We show in this study that testing of patients with known risk factors does not appear to be commonly accepted in practice, as our Acquired Pathology and Non-specific, Likely Causal groups had the lowest rates of genetic testing. Testing of our Normal and Congenital/Structural groups had higher rates of genetic testing, consistent with previous publications^16^. However, several studies found evidence to suggest that genetic diagnoses are still common in patients with established risk factors for CP. One study hypothesized that patients with certain risk factors such as extremely preterm birth, brain bleed or stroke, birth asphyxia, brain malformations, and intrauterine infection would have lower rates of genetic diagnosis than those without certain risk factors. Surprisingly, they were not able to prove any significant difference in genetic diagnosis rates^12^. In another study of 150 clinically heterogeneous patients with CP who underwent genome sequencing, 24.7% of patients carried a likely pathogenic or pathogenic variant, and an additional 34.7% carried a highly suspicious variant of unknown significance. Importantly, roughly half the pathogenic or likely pathogenic variants would change management of the patient’s disease process^13^. One group performed whole exome sequencing on a cohort of patients with CP and found a significant prevalence of Mendelian disorders in both cryptogenic and non-cryptogenic CP patients^19^. In a recent review of genetic findings in patients with CP, the authors argue that even patients with established risk factors may warrant genetic testing, arguing that patients with known gray or white matter injuries may have a genetic predisposition for such injuries^16^. Overall, our findings suggest that clinical decision making regarding genetic testing in CP may be determined based on brain MRI, despite evidence that suggests even patients with injury patterns may have an underlying genetic diagnosis. The de facto guideline for genetic testing in the setting of mild-MRI findings may be due to a lack of clear guidelines for CP genetic testing that takes into account MRI findings and gestational age.

Our study had several limitations. Our sample was limited in size due to drawing patients from a single tertiary care institute, as well as the limited availability of brain MRI reports. Due to the small sample size there was not enough power in our study to draw firm conclusions regarding the difference in CP and global disease severity, nor could we conclude whether there were differences in genetic testing and diagnostic yield between MRI categorization groups. Despite these limitations, the rates of all comorbidities, CP disease severity and genetic testing findings were similar across the groups. A final limitation of our study is that the most commonly accepted method of grading the severity of CP via the GMFCS categorization system was not used in the study due to lack of recorded scores in the electronic medical record.

## VI. Conclusion

To our knowledge, this is the largest descriptive study of a sample of children born between 32w0d and 33w6d with CP. In our sample, children with CP who were born between 32 and 33w6d commonly present with Acquired Pathology findings on brain MRI. Few patients had Normal MRI findings, suggesting that an abnormal MRI at these gestational ages is highly associated with CP. Surprisingly, disease comorbidities such as epilepsy, tracheostomy, and gastrostomy tube dependence did not vary significantly between MRI groups. Similarly, disease severity by CP markers as measured by verbal status, ambulatory status, and intellectual disability did not vary significantly between MRI groups. Genetic testing was utilized more often when imaging did not indicate a brain injury, but genetic testing still yielded a diagnosis 25% of the time.Given this consistency of disease severity across MRI groups, it is possible that additional factors such as genetic findings, may contribute to disease beyond brain injury. There may be a role for more genetic testing in this population. Further work is required to determine the kind of brain injuries most common in moderately preterm infants with CP. Future studies may also investigate the relationship between severity of brain MRI findings and clinical status of the patient with CP. In addition, further work must be done to elucidate the utility of genetic testing in patients with CP.

## Data Availability

All data produced in the present study are available upon reasonable request to the authors

## VIII. Acknowledgments

The authors would like to thank the Ann & Robert H Lurie Children’s Hospital of Chicago for their support in completing this project.

## IX. Author Contributions

Elizabeth Fisher – first authorship, preliminary literature review, study design, chart review, data analysis, manuscript editing

Jessica Tartakovsy – preliminary literature review, chart review, data analysis Laura A. Bliss MD – preliminary literature review, study design

Alok Jaju MD – study design

Jessie Aw-Zoretic MD – study design

Laura Vernon MD – study design

Divakar S. Mithal MD PhD – senior authorship, study design, data analysis, manuscript editing

## X. Statements and Declarations

### Ethical Considerations

This study received Institutional Review Board approval from the Ann & Robert H Lurie Children’s Chicago.

### Consent to Participate

Not applicable

### Consent for Publication

Not applicable

### Declaration of Conflicting Interest

The authors declared no potential conflicts of interest with respect to the research, authorship, and/or publication of this article.

### Funding

The authors received no financial support for the research, authorship, and/or publication of this article.

## Notes

### Competing Interest Statement

The authors have declared no competing interest.

### Funding Statement

This study did not receive any funding

### Author Declarations

The IRB of Ann and Robert H Lurie Children's Hospital of Chicago gave ethical approval for this work

